# Real-life evaluation of a rapid antigen test (Panbio™ COVID-19 Ag Rapid Test Device) for SARS-CoV-2 detection in asymptomatic close contacts of COVID-19 patients

**DOI:** 10.1101/2020.12.01.20241562

**Authors:** Ignacio Torres, Sandrine Poujois, Eliseo Albert, Javier Colomina, David Navarro

**Affiliations:** Microbiology Service, Hospital Clínico Universitario, INCLIVA Research Institute, Valencia, Spain; Department of Microbiology, School of Medicine, University of Valencia, Valencia, Spain

**Keywords:** SARS-CoV-2, COVID-19, rapid antigen detection test (RAD), asymptomatic, close contacts

## Abstract

**Objectives:** There is limited information on the performance of rapid antigen detection (RAD) tests to identify SARS-CoV-2-infected asymptomatic individuals. In this field study, we evaluated the Panbio™ COVID-19 Ag Rapid Test Device (Abbott Diagnostics, Jena, Germany) for the purpose.

**Methods:** A total of 634 individuals (355 female; median age, 37 years; range, 9-87) were enrolled. Household (n=338) contacts were tested at a median of 2 days (range, 1-7) after diagnosis of the index case and non-household contacts (n=296) at a median of 6 days (range, 1-7) after exposure. RAD testing was carried out at the point of care. The RT-PCR test used was the TaqPath COVID-19 Combo Kit (Thermo Fisher Scientific, Massachusetts, USA).

**Results:** In total, 79 individuals (12.4%) tested positive by RT-PCR, of whom 38 (48.1%) yielded positive RAD results. The overall sensitivity and specificity of the RAD test was 48.1% (95% CI: 37.4-58.9) and 100% (95% CI: 99.3-100), respectively. Sensitivity was higher in household (50.8%; 95% CI: 38.9-62.5) than in non-household (35.7%; 95% CI:16.3-61.2%) contacts. Individuals testing positive by RAD test were more likely (*P*<0.001) to become symptomatic than their negative counterparts.

**Conclusion:** The Panbio test displays low sensitivity in asymptomatic close contacts of COVID-19 patients, particularly in non-household contacts. Nonetheless, establishing the optimal timing for upper respiratory tract collection in this group seems imperative to pinpoint test sensitivity.

## INTRODUCTION

Rapid antigen detection (RAD) immunoassays have emerged as a valuable alternative to RT-PCR for diagnosis of SARS-CoV-2 infection in patients presenting with clinically compatible COVID-19 [1]. RAD tests are simple to carry out and return results within a short time, thus being well-suited for point-of-care testing (POCT). Moreover, RAD tests can be used as a proxy for SARS-CoV-2 cultured from respiratory tract specimens, thus allowing reasonably accurate prediction of contagiousness [2,3]. The possibility of using RAD tests to identify SARS-CoV-2-infected asymptomatic contacts of COVID-19 patients is appealing, as it could effectively contribute to minimize community SARS-CoV-2 spread through early detection of highly infectious individuals [1], yet little is known about how RAD tests perform in this population group [4-6]. Here, we report on the performance of the Panbio™ COVID-19 Ag Rapid Test Device (Abbott Diagnostic GmbH, Jena, Germany) conducted at POC in this setting.

## Material and methods

### Patients

A total of 634 consecutive asymptomatic individuals (female, n=355; median age, 37 years; range, 9-87 years) attended at the Clínico-Malvarrosa Health Department (Valencia, Spain) were enrolled between October 16 and November 20, 2020. Participants were either household (n=338) or non-household (n=296) close contacts of COVID-19 patients, as defined by the Spanish Ministry of Health [7]. Timing of sample collection was prescribed at the discretion of either the physician in charge of the index case or local health authorities. The study was approved by the Hospital Clínico de Valencia (HCU) INCLIVA Research Ethics Committee.

### SARS-CoV-2 testing

Nasopharyngeal swabs (NP) for RAD and RT-PCR testing were collected by experienced nurses at the POC site located at Hospital Malvarrosa, as previously detailed [3]. RAD testing was carried out at POC immediately after sampling. RT-PCRs were conducted within 24 h. of specimen collection at the Microbiology Service of Hospital Clínico Universitario (Valencia, Spain) with the TaqPath COVID-19 Combo Kit (Thermo Fisher Scientific, Massachusetts, USA). RT-PCR Ct values were normalized to copies/ml as previously described [3].

### Statistical analyses

Agreement between RAD and RT-PCR tests was assessed using Cohen’s Kappa (κ) statistics. Differences between medians were compared using the Mann-Whitney U-test. The Chi-squared test was used for frequency comparisons. Two-sided *P*-values <0.05 were considered statistically significant. Statistical analyses were performed using SPSS version 25.0 (SPSS, Chicago, IL, USA).

## Results

### Overall performance of the RAD test in asymptomatic close contacts

A total of 79 out of 634 individuals (12.4%) tested positive by RT-PCR, of whom 38 (48.1%) returned positive RAD test results. There were no RT-PCR positive/RAD negative cases. Accordingly, concordance between RT-PCR and RAD results was moderate (κ index, 0.61; 95% CI, 0.5-0.73). As shown in Figure 1, SARS-CoV-2 RNA load in NP was significantly higher (*P*<0.001) in RAD-positive (median, 8.7 log_10_ copies/ml) than in RAD-negative individuals (4.9 log_10_ copies/ml).

**Figure 1.**
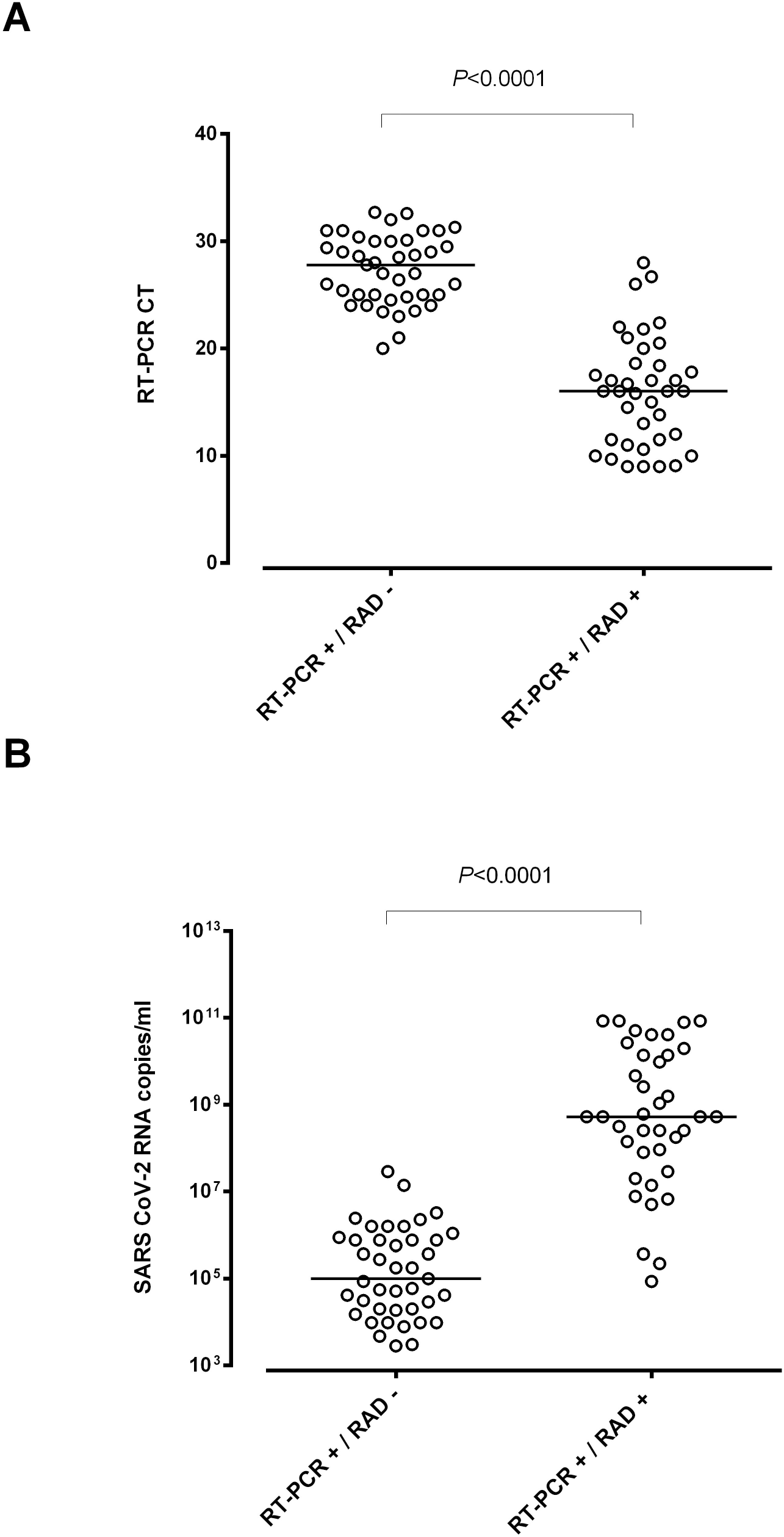
RT-PCR cycle thresholds (Ct) (A) and SARS-CoV-2 RNA load (B) in asymptomatic close contacts of COVID-19 patients testing either positive or negative by Panbio™ COVID-19 Ag Rapid Test Device (RAD). The AMPLIRUN® TOTAL SARS-CoV-2 Control (Vircell S.A:, Granada, Spain) was used as the reference material for SARS-CoV-2 RNA load quantitation (in copies/ml, considering RT-PCR Cts for the N gene [3]). *P* values for comparisons are shown.

Overall sensitivity and specificity of RAD was 48.1% and 100% (Table 1). For the above-mentioned prevalence (12.4%), the negative predictive value (NPV) of the RAD test was 94.5%. As expected, RAD sensitivity was directly related to SARS-CoV-2 load in NP specimens (Supplementary Table 1), reaching 96.8% when specimens with viral load ≥ 7.4 log10 copies/ml (Ct ≤20) were analyzed separately.

**TABLE 1.** Performance of the Panbio™ COVID-19 Ag Rapid Test Device for SARS-CoV-2 detection in asymptomatic household and non-household close contacts.

| Parameter | Population group |  |  |
| --- | --- | --- | --- |
|  | All individuals | Household contacts | Non-household contacts |
| Sensitivity % (95% CI) | 48.1 (37.4-58.9) | 50.8 (38.9-62.5) | 35.7 (16.3-61.2) |
| Specificity% (95% CI) | 100 (99.3-100) | 100 (98.6-100) | 100 (98.7-100) |
| Negative predictive value | 93.1 (90.8-94.9) | 89.5 (85.9-92.5) | 96.9 (94.2-98.4) |
| Positive predictive value | 100 (90.8-100) | 100 (89.6-100) | 100 (56.6-100) |

<sup>a</sup>Adjusted to actual prevalence in the respective population group.

### Performance of RAD test in household and non-household asymptomatic close contacts

Household contacts (n=338; median age, 36.5; range, 10-86 years; 175 female) were tested at a median of 2 days (range, 1-7) after diagnosis of the presumed index case. Sixty-five (19.2%) tested positive by RT-PCR, of whom 33 (50.7%) were positive by RAD test. The likelihood of obtaining either a positive or a negative RAD result was unrelated to the time elapsed since diagnosis of the index case (*P*=0.33).

Non-household contacts (n=296; median age, 38.5 years; range, 9-87 years; 180 female) were tested at a median of 6 days (range, 1-7) after self-reported exposure. Five individuals yielded RT-PCR-positive/RAD-positive results (1.6%) and 9 had RT-PCR-positive/RAD-negative results (3.0%). Overall, median time from exposure to testing was similar among individuals displaying either positive or negative RAD results (*P*=0.89).

The agreement level between RT-PCR and RAD results was significantly higher (*P*<0.001) for household (κ, 0.61; 95% CI, 0.50-0.75) than for non-household (κ, 0.51; 95% CI, 0.20-0.83) contacts. RAD sensitivity was significantly higher (*P* <0.001) in household contacts, while the opposite was true for NPV (Table 1).

SARS-CoV-2 RNA load was comparable (*P*=0.21) across household (median, 6.8 log_10_ copies/ml; range, 3.4-10.9) and non-household (median, 5.9 log_10_ copies/ml; range, 3.5-10.6) contacts, and was significantly higher (*P*<0.001) in RAD-positive than in RAD-negative individuals, irrespective of the subcohort considered.

### Clinical outcomes

Thirty-nine out of the 79 individuals testing positive by RT-PCR eventually became mildly symptomatic (49.3%), without requiring hospitalization. Individuals testing positive by RAD were more likely (*P*<0.001) to develop COVID-19 (30 out of 38) than those who did not (9 out of 41).

## Discussion

In this field study, overall sensitivity of the Panbio™ COVID-19 Ag Rapid Test Device for identification of SARS-CoV-2-infected individuals among asymptomatic close contacts of confirmed COVID-19 cases was 48.1%, close to the figures reported by Linares et al. (54.5%) [4], Fenollar et al. (45.4%) [5] and Bulilete et al. (59.0%) [6], in apparently comparable cohorts. However, in two of these studies [4,5], the RAD test was carried out at a central laboratory, and timing of sample collection was not disclosed [4,5]. In the study by Bulilete at al. [5] most participants (70.6%) were tested within 5 days of exposure. Sensitivity of the Panbio^™^ test was lower than was previously found [3-6] in symptomatic patients (around 80%), yet as reported for the latter patients, RAD sensitivity was directly related to the magnitude of SARS-CoV-2 RNA load in NP specimens. Such a striking difference might reflect dissimilarities across symptomatic and asymptomatic individuals in the kinetics of SARS-CoV-2 load in the upper respiratory tract [8,9]. While it is well known that SARS-CoV-2 load peaks around the time of symptoms onset in the former group [10,11], the timing is uncertain in asymptomatic cases.

Interestingly, individuals testing positive by RAD were more likely to become (mildly) symptomatic than their negative counterparts, pointing to a pathogenetic link between SARS-CoV-2 RNA load and development of overt COVID-19.

The strength of the current study is that it reflects the real-life performance of the RAD test at POC. Among its limitations are the relative low number of cases, and the possibility that samples were collected too early after exposure, particularly in non-household contacts, in whom RAD sensitivity was strikingly low. In this sense, Linares et al. [4] reported the sensitivity of the Panbio^™^ test as very low in close contacts at less than 7 days from exposure.

In summary, we found the Panbio™ test to display low sensitivity in asymptomatic contacts of COVID-19 patients. Nevertheless, establishing the optimal timeframe for NP collection in household and non-household contacts seems crucial to accurately determine the sensitivity of the test.

## Data Availability

The authors confirm that the data supporting the findings of this study are available within the article [and/or] its supplementary materials.

## ACKNOWLEDGMENTS

We are grateful to Abbott Diagnostics for providing the Panbio™ COVID-19 Ag Rapid Test Device kits. We thank all personnel working at Clinic University Hospital and primary healthcare centers belonging to the Clínico Malvarrosa Health Department for their unwavering commitment in the fight against COVID-19. We would also like to thank María José Beltrán, Pilar Botija and Ana Sanmartín for assistance in organizing RAD testing in primary healthcare centers and Salvador Peiró for critical revision of the manuscript.

## AUTHOR’S CONTRIBUTIONS

IT, SP and EA: Methodology and data validation. EA, IT and JC: Formal analysis. DN: Conceptualization, supervision, writing the original draft. All authors reviewed the original draft.

