## Supplementary Table 1 for "Real-life evaluation of a rapid antigen test (Panbio™ COVID-19 Ag Rapid Test Device) for SARS-CoV-2 detection in asymptomatic close contacts of COVID-19 patients"

| **SUPPLEMENTARY TABLE 1. Overall sensitivity of the Panbio™ COVID-19 Ag Rapid Test Device according to the SARS-CoV-2 RNA load in nasopharyngeal specimens** | | |
| --- | --- | --- |
| RT-PCR cycle threshold value | SARS-CoV-2 RNA load (log_10_ copies/ml) | Sensitivity (95% CI) |
| ≤ 20 | ≥7.4 | 96.8 (83.8-99.4) |
| ≤ 25 | ≥5.8 | 71.4 (57.6-82.2) |
| ≤ 30 | ≥4.3 | 55.1 (43.4-66.2) |
| ≤ 35 | ≥2.7 | 48.1 (37.4-58.9) |
